# Immunogenicity and Safety Following a Homologous Booster Dose of a SARS-CoV-2 recombinant spike protein vaccine (NVX-CoV2373): A Phase 2 Randomized Placebo-Controlled Trial

**DOI:** 10.1101/2021.12.23.21267374

**Authors:** Raburn Mallory, Neil Formica, Susan Pfeiffer, Bethanie Wilkinson, Alex Marcheschi, Gary Albert, Heather McFall, Michelle Robinson, Joyce S. Plested, Mingzhu Zhu, Shane Cloney-Clark, Bin Zhou, Gordon Chau, Andreana Robertson, Sonia Maciejewski, Gale Smith, Nita Patel, Gregory M. Glenn, Filip Dubovsky, for the Novavax Inc. 2019nCoV-101 Study Group

## Abstract

**Background:** Emerging SARS-CoV-2 variants and evidence of waning vaccine efficacy present significant obstacles toward controlling the COVID-19 pandemic. Booster doses of SARS-CoV-2 vaccines may address these concerns by both amplifying and broadening the immune responses seen with initial vaccination regimens.

**Methods:** In a phase 2 study, a single booster dose of a SARS-CoV-2 recombinant spike protein vaccine with Matrix-M™ adjuvant (NVX-CoV2373) was administered to healthy adult participants 18 to 84 years of age approximately 6 months following their primary two-dose vaccination series. Safety and immunogenicity parameters were assessed, including assays for IgG, MN_50_, and hACE2 receptor binding inhibition against the ancestral SARS-CoV-2 strain and select variants (B.1.351 [Beta], B.1.1.7 [Alpha], B.1.617.2 [Delta], and B.1.1.529 [Omicron]). This trial is registered with ClinicalTrials.gov, NCT04368988.

**Findings:** An incremental increase in the incidence of solicited local and systemic reactogenicity events was observed with subsequent vaccinations. Following the booster, incidence rates of local and systemic reactions were 82.5% (13.4% ≥ Grade 3) and 76.5% (15.3% ≥ Grade 3), respectively, compared to 70.0% (5.2% ≥ Grade 3) and 52.8% (5.6% ≥ Grade 3), respectively, following the primary vaccination series. Events were primarily mild or moderate in severity and transient in nature, with a median duration of 1.0 to 2.5 days. Immune responses seen 14 days following the primary vaccination series were compared with those observed 28 days following the booster (Day 35 and Day 217, respectively). For the ancestral SARS-CoV-2 strain, serum IgG geometric mean titers (GMTs) increased ∼4.7-fold from 43,905 ELISA units (EU) at day 35 to 204,367 EU at Day 217. Neutralization (MN_50_) assay GMTs showed a similar increase of ∼4.1-fold from 1,470 at day 35 to 6,023 at Day 217. A functional hACE2 receptor binding inhibition assay analyzing activity against ancestral and variant strains of SARS-CoV-2 at Day 189 vs Day 217 found 54.4-fold (Ancestral), 21.9-fold (Alpha), 24.5-fold (Beta), 24.4-fold (Delta), and 20.1-fold (Omicron) increases in titers. An anti-rS IgG activity assay comparing the same time points across the same SARS-CoV-2 strains found titers improved 61.2-fold, 85.9-fold, 65.0-fold, 92.5-fold, and 73.5-fold, respectively.

**Interpretation:** Administration of a booster dose of NVX-CoV2373 approximately 6 months following the primary vaccination series resulted in an incremental increase in reactogenicity along with enhanced immune responses. For both the prototype strain and all variants evaluated, immune responses following the booster were notably higher than those associated with high levels of efficacy in phase 3 studies of the vaccine.

**Funding:** Novavax® and the Coalition for Epidemic Preparedness Innovations (CEPI®).

## INTRODUCTION

The coronavirus disease 2019 (COVID-19) pandemic began shortly after the emergence of the novel severe acute respiratory syndrome coronavirus 2 (SARS-CoV-2) in Wuhan, China in December of 2019. Since then, SARS-CoV-2 transmission has continued, resulting in millions of deaths globally due to COVID-19. Multiple vaccines produced using various technologies have been developed to confer immunity against the ancestral SARS-CoV-2 spike (S) protein (mRNA-1273; BNT162b2; AD26.COV2.S; AZD1222), with approximately 8 billion doses administered to date (Ritchie 2021). Although clinical studies and real-world evidence have shown that these vaccines are highly effective in reducing severe disease and death due to COVID-19, there are also signs that their protection may be waning since vaccination campaigns began in late 2020. In addition, several SARS-CoV-2 variants have emerged with changes in the S protein receptor binding domain (RBD), enabling them to partially avoid neutralization in vaccinated persons. Five such variants have been classified by the World Health Organization (WHO) as variants of concern (VOCs): B.1.1.7 (Alpha), B.1.351 (Beta), P.1 (Gamma), B.1.617.2 (Delta), and most recently B.1.1.529 (Omicron). The development of variant-specific vaccines and the use of homologous vaccine booster doses are both being investigated as potential avenues to bolster protection against SARS-CoV-2 and its variants.

Novavax has developed a SARS-CoV-2 recombinant S protein nanoparticle vaccine (SARS-CoV-2 rS), comprising the full length, pre-fusion trimers of the ancestral SARS-CoV-2 S glycoprotein, co-formulated with a saponin-based adjuvant, Matrix-M™ (NVX-CoV2373). Favorable vaccine efficacy (VE) for NVX-CoV2373 against mild, moderate, or severe COVID-19 was established in two phase 3, randomized, placebo-controlled clinical trials of healthy and medically stable adult participants using a two-dose series (two doses administered 3 weeks apart). One trial of 15,139 participants in the United Kingdom (UK) found VEs of 89.7% (95% CI: 80.2, 94.6) for all participants, 86.3% (95% CI: 71.3, 93.5) for the B.1.1.7 (Alpha) variant and 96.4% (95% CI: 73.8, 99.5) for the ancestral strain. In a second trial of 29,582 participants in the United States of America (USA) and Mexico, the resultant VE was 90.4% (95% CI: 82.9, 94.6) for all participants, 100.0% (95% CI: 80.8, 100.0) for non-VOC or variant of interest (VOI), 93.2% (95% CI: 83.9, 97.1) for VOC/VOIs, and 93.6% (81.7, 97.8) for participants whose SARS-CoV-2 variant was B.1.1.7 (Alpha) (Dunkle 2021, Heath 2021).

As part of an ongoing phase 2, randomized, placebo-controlled clinical trial of NVX-CoV2373, a single booster dose was administered to healthy adult participants approximately 6 months following their primary two-dose vaccination series (Formica 2021). Immunogenicity and safety data through 28 days following the booster dose are now available and can be compared with those observed following the primary vaccination series in the same participant population.

## METHODS

### Study Design

A single booster dose was administered to participants as part of an ongoing phase 2, randomized, observer-blinded, placebo-controlled study of NVX-CoV2373.

NVX-CoV2373 was developed by Novavax®, who designed and sponsored the study, with funding support from the Coalition for Epidemic Preparedness Innovations (CEPI). The trial protocol was approved by the Alfred Hospital Ethics Committee (Melbourne, Victoria, Australia) and Advarra Central Institutional Review Board (Colombia, Maryland, USA) and is registered on Clinicaltrials.gov (NCT04368988).

### Participants

Healthy male and female participants ≥ 18 to ≤ 84 years of age were recruited for enrollment in this study.

Participants were eligible if they had a body mass index of 17 to 35 kg/m^2^, were able to provide informed consent prior to enrollment, and (for female participants) agreed to remain heterosexually inactive or use approved forms of contraception.

Participants with a history of severe acute respiratory syndrome (SARS) or a confirmed diagnosis of COVID-19, serious chronic medical conditions (eg, diabetes mellitus, congestive heart failure, autoimmune conditions, malignancy), or that were currently being assessed for an undiagnosed illness which may lead to a new diagnosis, were excluded from the study. Pregnant or breastfeeding females were also excluded.

Written informed consent was obtained from all participants prior to their enrollment.

### Randomization and Masking

The randomization schemes used in this trial were executed using a centralized interactive response technology (IRT) system according to pre-generated randomization schedules. At the start of the trial, participants were randomly assigned to one of five initial treatment groups (1:1:1:1:1) as previously described by Formica (Formica 2021; Figure 1). Of the five treatment groups, one was a placebo control (Group A) and two were active vaccine groups that were considered for additional vaccination with a booster (Group B and Group C). After approximately 6 months, consenting participants who had been randomized to receive a primary vaccination series of either two doses of NVX-CoV2373 (5 μg SARS-CoV-2 rS + 50 μg Matrix-M adjuvant) on Day 0 and Day 21 (Group B) or one dose of NVX-CoV2373 (5 μg SARS-CoV-2 rS + 50 μg Matrix-M adjuvant) on Day 0 and placebo on Day 21 (Group C) were re-randomized 1:1 to receive either a single booster dose of NVX-CoV2373 at the same dose level (Groups B2 and C2) or placebo (Groups B1 or C1) at Day 189. Participants could decline re-randomization for any reason (eg, they had already received an emergency authorized vaccine). Group B participants are the main focus of this article as they were vaccinated using the NVX-CoV2373 dose and regimen proposed for approval and commercial distribution. All other treatment groups are continuing to be followed for safety events only.

**Figure 1.**
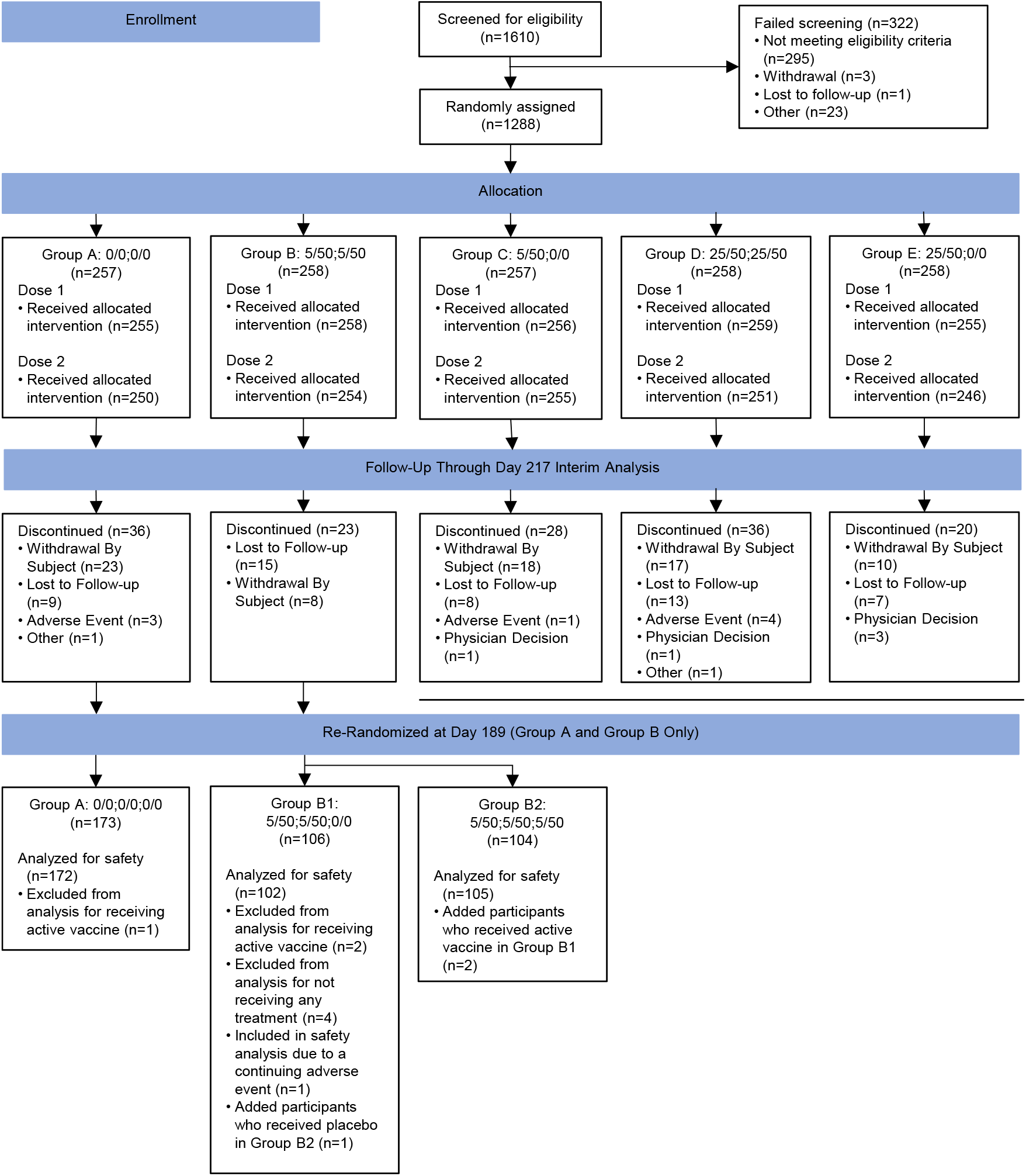
Consort Diagram for Study 2019nCoV-101 (Part 2): Booster Dosing of Group B Participants.

All vaccinations were prepared and administered on an outpatient basis by designated site personnel in a way to maintain the blind. Participants and other site staff remained blinded to individual treatment assignment.

### Procedures

All active booster vaccinations were administered at the same dose level as the primary vaccination series (5 µg SARS-CoV-2 rS with 50 µg Matrix-M adjuvant) via intramuscular (IM) injection. All booster vaccinations, active or placebo, were administered at an injection volume of 0.5 mL.

Participants utilized an electronic diary to record reactogenicity on the day of vaccination and for an additional 6 days thereafter. Blood samples for immunogenicity analysis were collected 28 days after receipt of the booster, with safety follow-up also being performed at this time.

### Assays

Measures of immune response included assays for serum immunoglobulin G (IgG) antibodies, neutralizing antibody activity (microneutralization assay at an inhibitory concentration >50% [MN_50_]), and human angiotensin-converting enzyme 2 (hACE2) receptor binding inhibition. Serum IgG antibody levels specific for the SARS-CoV-2 rS protein antigen were detected using a qualified or validated IgG ELISA (Novavax Clinical Immunology, Gaithersburg, MD, US) for the ancestral strain and Beta variant, respectively. Neutralizing antibodies specific for SARS-CoV-2 virus were measured using a qualified or validated wild-type virus (Victoria strain or Beta variant) MN_50_ assay (360biolabs Melbourne Vic Australia) for the ancestral strain and Beta variant, respectively. Serum IgG and MN_50_ assay data were collected for both the ancestral and Beta variant SARS-CoV-2 strains. Fit-for-purpose functional hACE2 receptor binding inhibition and an anti-rS (anti-recombinant spike) IgG activity assays were both used to analyze responses against the ancestral, B.1.351 (Beta), B.1.1.7 (Alpha), B.1.617.2 (Delta), and B1.1.529 (Omicron) variant strains of SARS-CoV-2.

### Outcomes

Safety outcomes included participant-reported reactogenicity events for 7 days following the booster, as well as unsolicited adverse events occurring through 28 days post-booster. Booster reactogenicity was documented separately by solicited local and systemic adverse events. Unsolicited adverse events from booster vaccination to 28 days post-booster were recorded. Data were also collected on whether an adverse event was serious, related to vaccination, related to COVID-19, a potentially immune-mediated medical condition (PIMMC), or lead to discontinuation or an unscheduled visit to a healthcare practitioner.

Participant samples for immunogenicity analyses were collected immediately prior to and 28 days after the booster.

### Statistical Analysis

Analyses included safety and immunogenicity data from participants in Group B obtained during and after their primary vaccination series (Day 0, Day 21, Day 35, Day 105, and Day 189) for comparison with data collected from Group B2 28 days following their receipt of the booster dose (Day 217). Results were also analyzed by participant age group: ≥ 18 to ≤ 84 years of age, ≥ 18 to ≤ 59 years of age, and ≥ 60 to ≤ 84 years of age.

The safety analysis included all participants who received a single booster injection of NVX-CoV2373 (Group B2) or placebo (Group B1). Safety analyses were presented as numbers and percentages of participants with solicited local and systemic adverse events analyzed through 7 days after each vaccination and unsolicited adverse events through 28 days following the booster.

The Per-Protocol (PP) Analysis Set was used for immunogenicity analysis. The PP Analysis Set was determined by study visit and included all participants who received the primary vaccination series on Day 0 and Day 21 (SARS-CoV-2 rS or placebo) for all analyses at Day 35, and all participants who received the primary vaccination series on Day 0 and Day 21 and the booster dose on Day 189 for all analyses at Day 217. All participants in the PP Analysis Set were analyzed according to the randomized treatment assignment. Any participant who was SARS-CoV-2 positive by qualitative PCR testing from screening and prior to the immunogenicity assessment for a particular time point was excluded from the PP Analysis Set for that time point onward.

## RESULTS

A total of 1610 participants were screened from 24 August 2020 to 25 September 2020, and 1,283 participants were enrolled and treated. All but three participants randomized to Group B (n=257) received both doses of NVX-CoV2373 in their primary vaccination series and were considered for investigation of a single booster dose at the same dose level (Figure 1). Re-randomization of Group B participants took place at Day 189, with 210 consenting participants assigned 1:1 to receive a single booster of NVX-CoV2373 in Group B2 (n=104) or placebo in Group B1 (n=106). In Group B2, all but one participant received active vaccine as a booster; this participant received placebo in error and was assessed for safety in Group B1. All but six participants in Group B1 received placebo as a booster; of the remaining six participants, four did not receive any booster (of which, one was included in Group B1 for safety due to an ongoing adverse event) and two received active vaccine in error as a booster and were assessed for safety in Group B2. All but one participant in Group A received placebo for all three doses, with the remaining participant receiving active vaccine as a booster dose.

Demographics and baseline characteristics were generally balanced between the active (Group B2) and placebo (Group B1) booster groups (Table 1), except for a higher proportion of female participants in Group B1 (58%) than Group B2 (45%). Across Groups A, B1, and B2, the median age was approximately 57 years and 45% of participants were ≥ 60 to ≤ 84years of age. Most participants were White (87%) and not Hispanic or Latino (95%). Baseline SARS-CoV-2 serostatus was predominantly negative (98%).

**Table 1.**
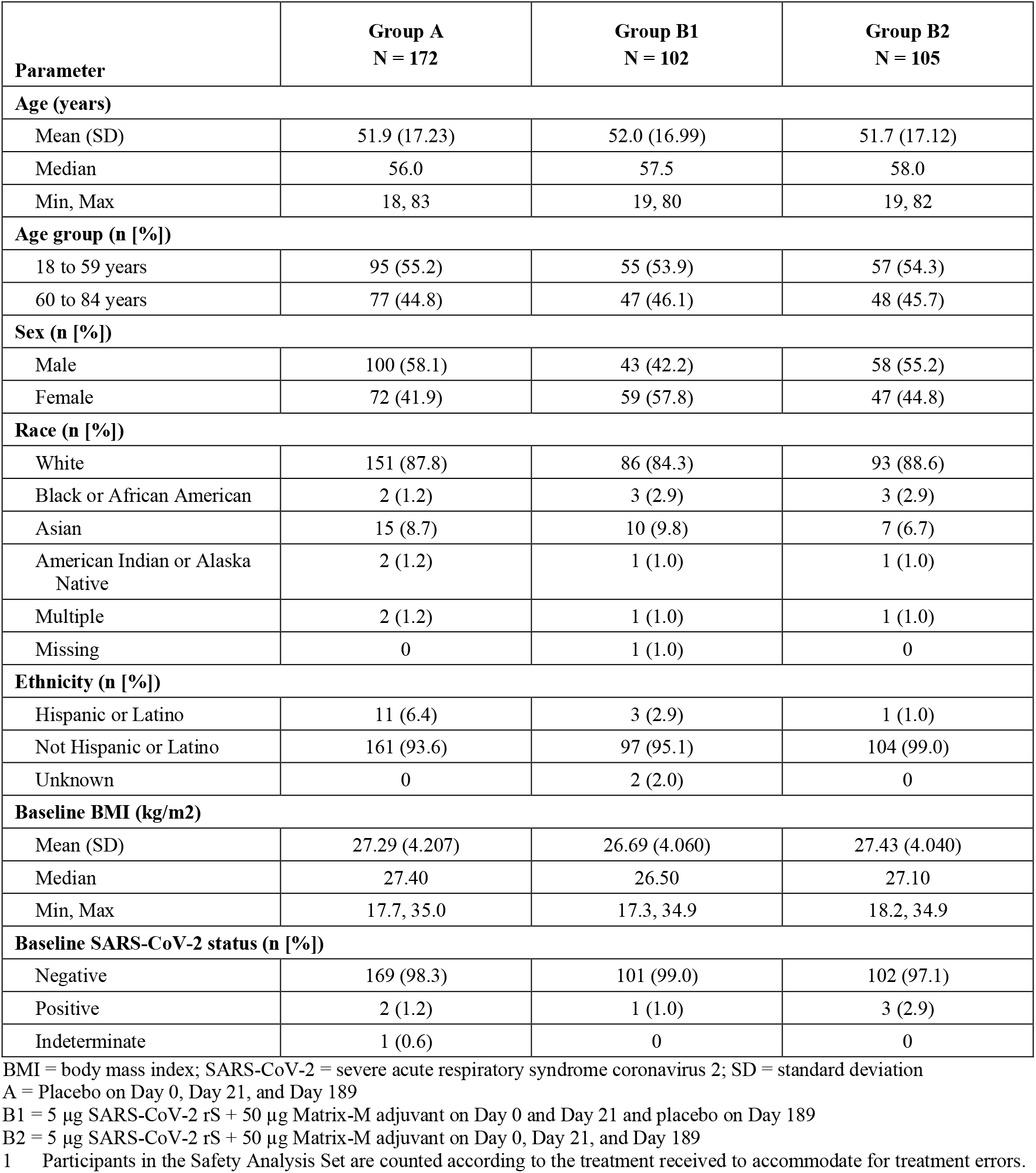
Demographics and Baseline Characteristics for Groups A, B1, and B2 (Safety Analysis Set^1^)

Safety reporting of solicited local and systemic reactogenicity events showed an increasing trend across all three doses of NVX-CoV2373 (Figure 2). Following the booster, participants in Group B2 reported an incidence rate for any local reaction (tenderness, pain, swelling, and erythema) of 82.5% (13.4% ≥ Grade 3) compared to 70.0% (5.2% ≥ Grade 3) following the primary vaccination series. Grade 4 local reactions were rare, with two events (pain and tenderness) reported by one participant in Group B2 compared with no participants following the primary vaccination series. Following the booster, local reactions were short-lived with a median duration of 2.0 days for all events except erythema (2.5 days). Local reactions were also short-lived following the primary vaccination series, with median durations of 2.0 days for pain and tenderness and 1.0 day for erythema and swelling.

**Figure 2.**
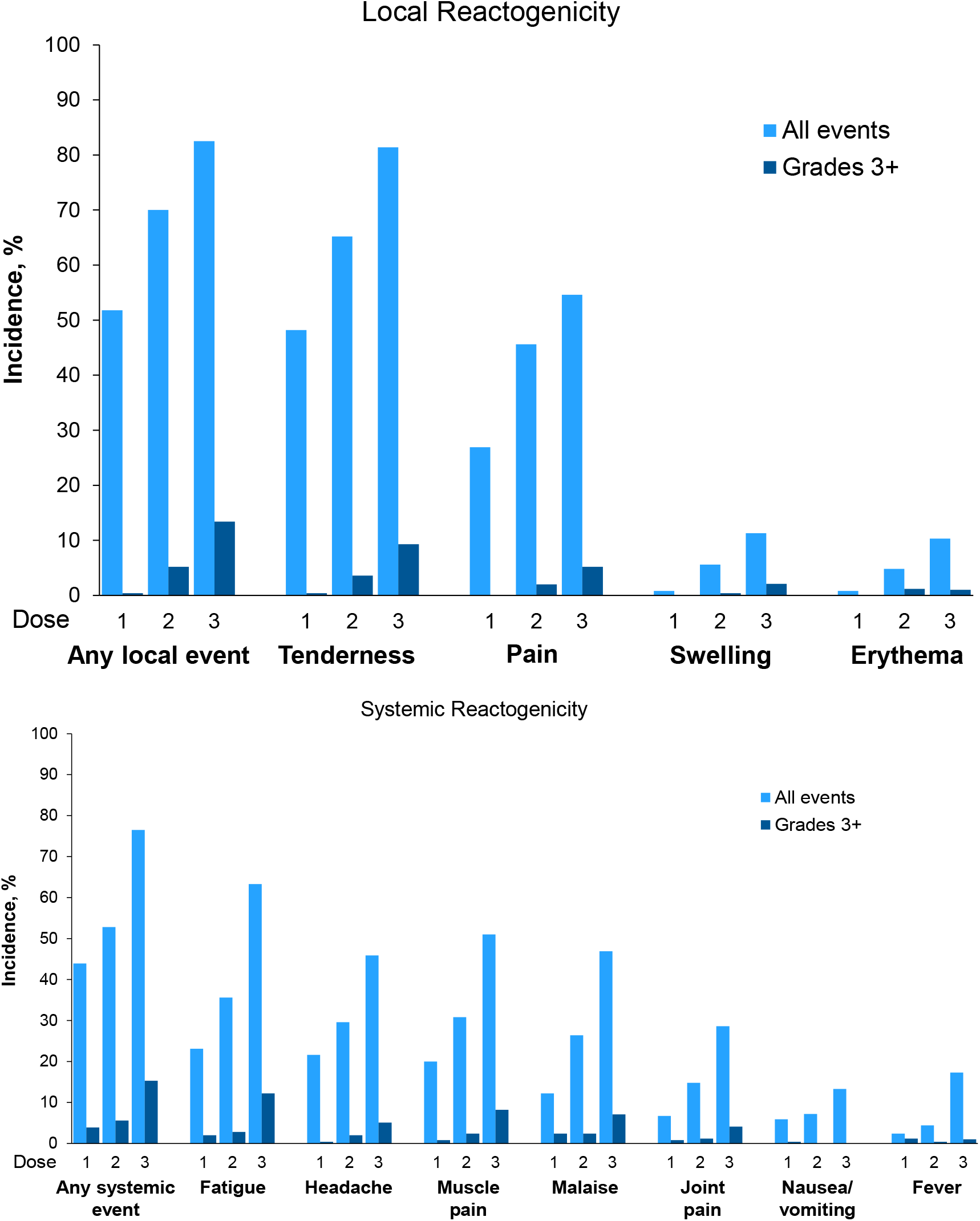
Local and Systemic Vaccine Reactogenicity by NVX-CoV2373 Dose (Group B2)

Systemic reactions showed a similar pattern with an incidence rate for any event (fatigue, headache, muscle pain, malaise, joint pain, nausea/vomiting, and fever) of 76.5% (15.3% ≥ Grade 3), compared to 52.8% (5.6% ≥ Grade 3), following the primary vaccination series. Grade 4 systemic reactions were rare, with three events reported by one participant in Group B2 (headache, malaise, and muscle pain) compared with no participants following the primary vaccination series. Following the booster, systemic reactions were transient in nature with a median duration of 1.0 day for all events except muscle pain which had a duration of 2.0 days. All systemic reactions were also short-lived following the primary vaccination series, with a median duration of 1.0 day for all events.

Local and systemic reactogenicity events were less frequent and less severe in older adults (≥ 60 to ≤ 84 years of age) when compared to younger adults (≥ 18 to ≤ 59 years of age) following either the primary vaccination series or booster dose. In the younger cohort, post-booster local and systemic reactions were reported in 84.9% (18.9% ≥ Grade 3) and 84.9% (26.4% ≥ Grade 3) of participants, respectively, versus 79.5% (6.8% ≥ Grade 3) and 66.7% (2.2% ≥ Grade 3) of participants, respectively, in the older cohort.

Unsolicited adverse events were summarized across the active-boosted participants (Group B2), placebo-boosted participants (Group B1), and participants receiving three doses of placebo throughout the study (Group A). Through 28 days after the booster, participants who initially received active vaccine for their primary vaccination series (Groups B2 and B1) experienced a higher incidence of unsolicited adverse events than those who received only placebo (Group A), with 12.4%, 12.7%, and 11.0% of participants reporting such events, respectively. A similar trend was seen for unsolicited severe adverse events (5.7%, 3.9%, and 2.4%, respectively). Other types of AEs reported included medically attended AEs (events requiring a healthcare visit; MAAEs), PIMMCs, events relevant to COVID-19, and serious adverse events (SAEs).

Overall, MAAEs occurred with a slightly higher frequency in active boosted participants across the three groups (30.5%, 26.1%, and 23.2% for Groups B2, B1, and A, respectively), with related events reported in few participants (1.9%, 0%, and 1.2%, respectively). Events considered PIMMCs were rare across the study, with one participant in Group B2 and Group A reporting a single event each; both events were assessed as not related to study treatment. No participant reported an as adverse event related to COVID-19.

SAEs were also infrequent across the study, occurring in 5.7%, 3.3%, and 1.6% of participants in Groups B2, B1, and A, respectively, with all events assessed as not related to study treatment.

Evaluation of SAEs for Group B2 and B1 participants did not show a relationship of with active boosting, as SAEs occurred in 0%, 4.8%, and 1.0% of participants in Group B2 and 0%, 2.0%, and 2.0% of participants in Group B1 following Dose 1, Dose 2, and the booster, respectively.

As expected, declines in Group B IgG and MN_50_ geometric mean titers (GMTs) were observed following the primary vaccination series (Day 35) through Day 189 (43,905 ELISA units [EU] to 6,064 EU for IgG and 1,470 to 63 for MN_50_, respectively). Twenty-eight days following the booster (Day 217), IgG and MN_50_ titers increased robustly compared to both the pre-booster titers and to the Day 35 titers produced by the primary series (Figure 3,Figure 4).

**Figure 3.**
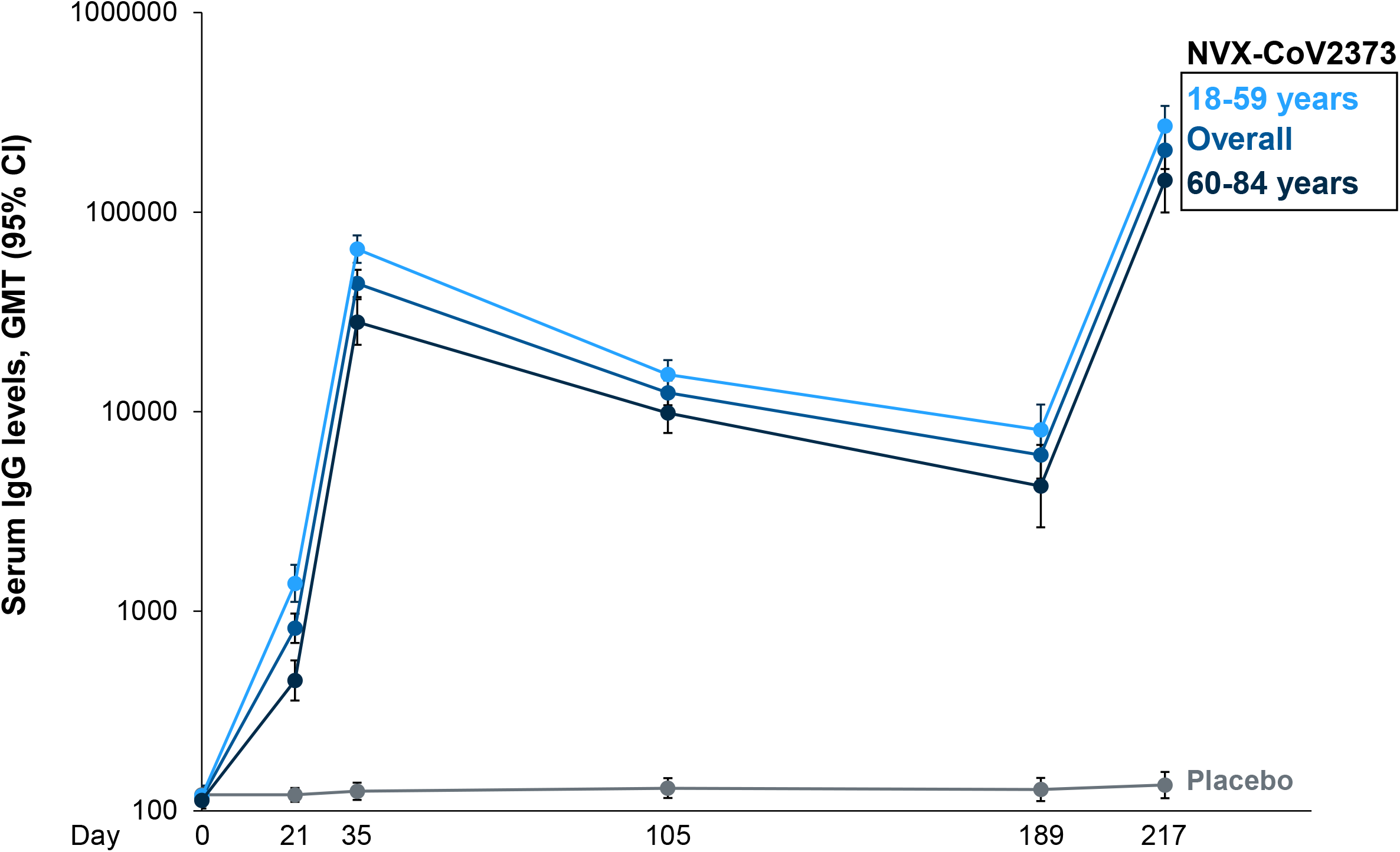
Serum IgG Titers to the Ancestral SARS-CoV-2 Strain by Study Day (Log Scale)

**Figure 4.**
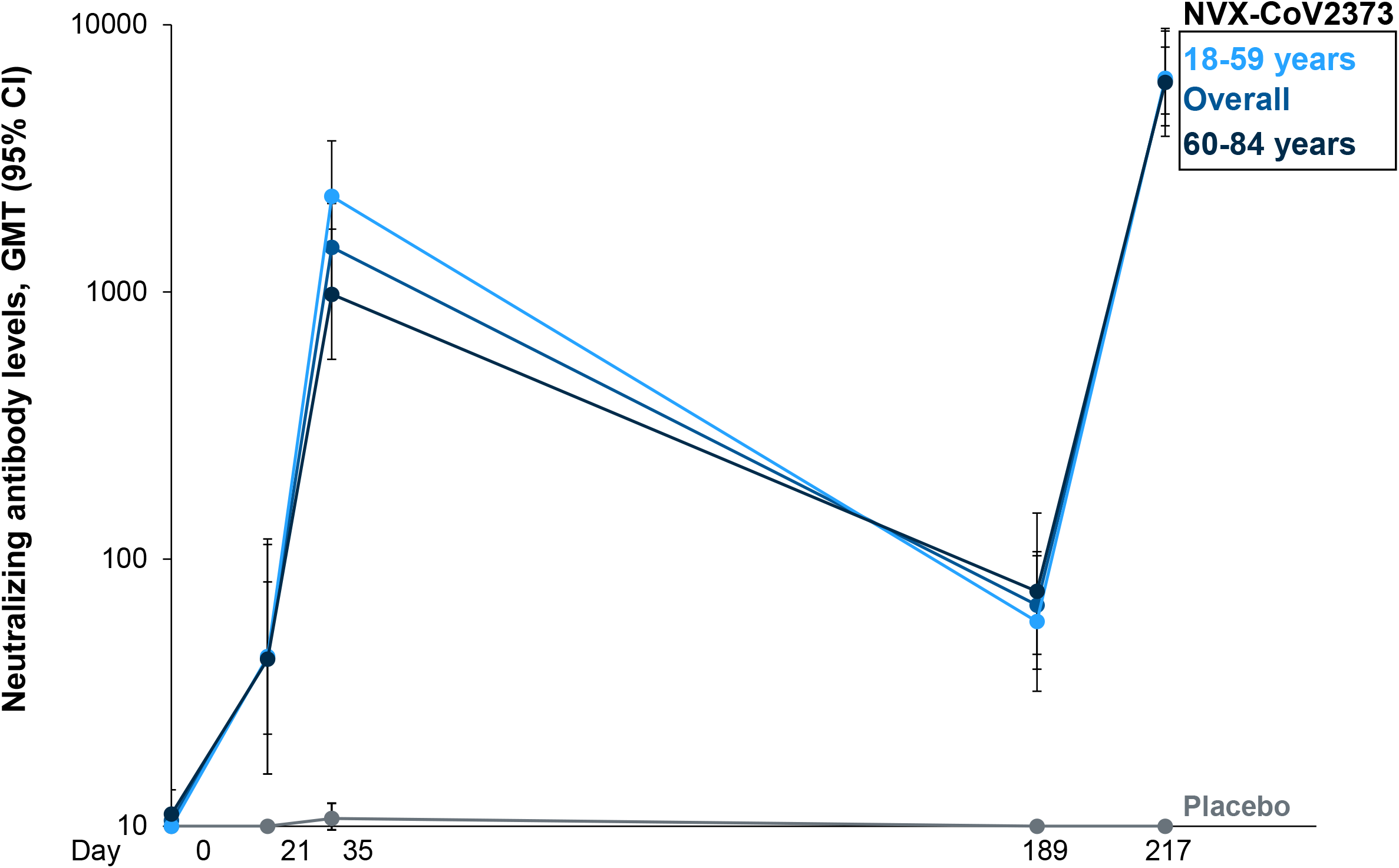
Neutralizing Antibody Activity for the Ancestral SARS-CoV-2 Strain by Study Day (Log Scale)

For the ancestral SARS-CoV-2 strain, serum IgG GMTs increased ∼4.7-fold from 43,905 EU following the primary vaccination series (Day 35) to 204,367 EU following the booster (Day 217). Higher fold increases after boosting were seen in older adults (5.1-fold) compared to younger adults (4.1-fold). Similarly, MN_50_ assay GMTs specific to the ancestral SARS-CoV-2 strain increased ∼4.1-fold from 1,470 to 6,023 over the same respective time points with increases in older and younger adults of 4.0-fold and 3.8-fold, respectively.

For the Beta variant, IgG GMTs increased from 4,317 EU at Day 189 pre-booster to 175,190 EU at Day 217 reflecting a post-booster increase of ∼40.6-fold. These titers were 4-fold higher than those observed at Day 35 for the ancestral strain (GMT 175,190 EU vs 43,905 EU). Beta variant MN_50_ assay data showed a similar fold increase in titers from pre-booster (Day 189) to post-booster (Day 217) of ∼50.1-fold (GMT 13 vs 661), though titers were lower than those seen for the ancestral strain at Day 35 (GMT 661 vs 1,470). (Table 2, Table 3).

**Table 2.** Serum IgG Geometric Mean Titers after Primary and Booster Vaccination for the.

| Age Group | Serum IgG GMT (EU [95% CI]) |  |  |  |  |
| --- | --- | --- | --- | --- | --- |
|  | Day 35<br>Ancestral Strain | Day 189<br>Ancestral Strain | Day 217<br>Ancestral Strain | Day 189<br>Beta variant | Day 217<br>Beta variant |
| All Participants,<br>18 to 84 years | 43,905<br>(37,500, 51,403) | 6,064<br>(4,625, 7,952) | 204,367<br>(164,543, 253,828) | 4,317<br>(3,261, 5,715) | 175,190<br>(139,895, 219,391) |
| Participants<br>18 to 59 years | 65,255<br>(55,747, 76,385) | 8,102<br>(6,041, 10,866) | 270,224<br>(214,304, 340,736) | 6,310<br>(4,642, 8,578) | 226,103<br>(176,090, 290,321) |
| Participants<br>60 to 84 years | 28,137<br>(21,617, 36,623) | 4,238<br>(2,631, 6,826) | 144,440<br>(99,617, 209,431) | 2,700<br>(1,682, 4,333) | 127,601<br>(86,809, 187,561) |

CI = confidence interval; ELISA = enzyme-linked immunosorbent assay; EU = ELISA unit; GMT = geometric mean titer; SARS-CoV-2 = severe acute respiratory syndrome coronavirus 2
Note: The ancestral strain IgG assay method is qualified, and the Beta variant IgG assay method is validated.

**Table 3.**
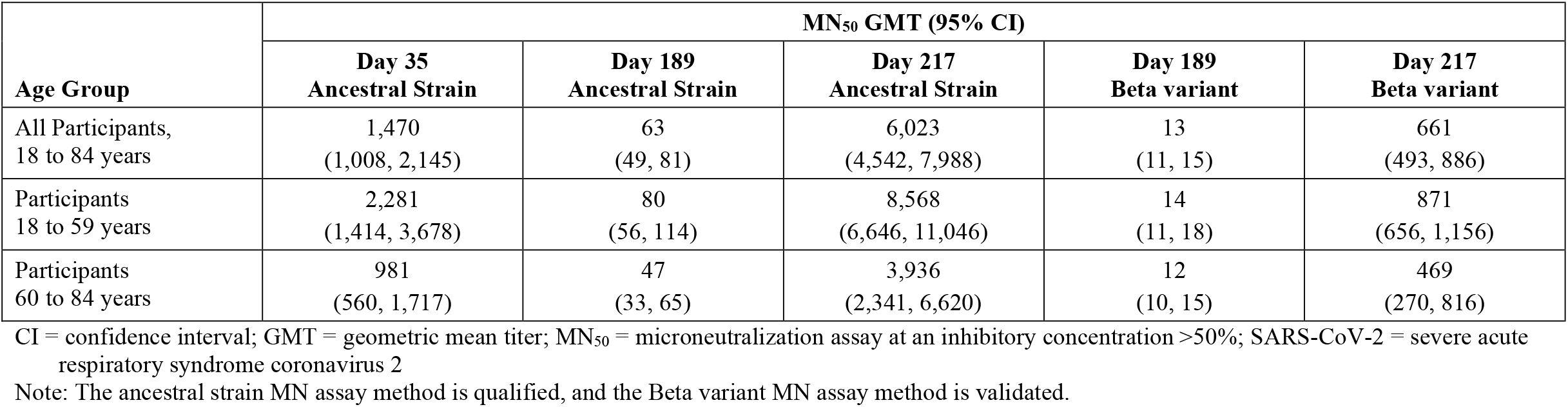
Neutralizing Antibody Activity after Primary and Booster Vaccination for the Ancestral and Beta Variant SARS-CoV-2 Strains by Study Day for Participants Receiving NVX-CoV2373.

Two assays were developed to assess immune responses against additional SARS-CoV-2 variants using participant sera from Group B2. A functional hACE2 receptor binding inhibition assay was utilized to compare activity against the ancestral strain and the Alpha, Beta, Delta, and Omicron variants of SARS-CoV-2 (Table 4, Figure 5). In respective order, 54.4-fold, 21.9-fold, 24.5-fold, 24.4-fold, and 20.1-fold increases in hACE2 inhibition titers were observed from Day 189 (immediately pre-booster) to Day 217. A second assay comparing anti-rS IgG activity across the same strains of SARS-CoV-2 found that 61.2-fold (Ancestral), 85.9-fold (Alpha), 65.0-fold (Beta), 92.5-fold (Delta), and 73.5-fold (Omicron) higher titers were observed after the booster (Table 5, Figure 6).

**Table 4.** hACE2 Receptor Binding Inhibition Geometric Mean Titers after Primary and Booster Vaccination for Ancestral and Variant SARS-CoV-2 Strains by Study Day for Participants Receiving NVX-CoV2373.

| Parameter | hACE2 Receptor Binding Inhibition Titers (IC<sub>50</sub>) |  |  |  |  |  |  |  |  |
| --- | --- | --- | --- | --- | --- | --- | --- | --- | --- |
| Strain | Ancestral |  |  | Alpha |  |  | Beta |  |  |
| Study Day | Day 35 | Day 189 | Day 217 | Day 35 | Day 189 | Day 217 | Day 35 | Day 189 | Day 217 |
| GMT<br>(95% CI) | 119.6<br>(78.7, 181.9) | 13.3<br>(10.0, 17.6) | 723.1<br>(533.5, 980.0) | 28.7<br>(20.0, 41.1) | 10.7<br>(9.3, 12.3) | 234.4<br>(170.2, 322.8) | 24.6<br>(16.7, 36.0) | 10.8<br>(9.18, 12.8) | 265.2<br>(189.3, 371.5) |
| GMFR D217/D35<br>(95% CI) | 6.1<br>(3.8, 9.9) |  |  | 8.1<br>(5.6, 11.9) |  |  | 10.8<br>(7.1, 16.4) |  |  |
| GMFR D217/D189<br>(95% CI) | 54.4<br>(37.0, 79.8) |  |  | 21.9<br>(15.07, 31.9) |  |  | 24.5<br>(16.5, 36.4) |  |  |
| Strain | Delta |  |  | Omicron |  |  |  |  |  |
| Study Day | Day 35 | Day 189 | Day 217 | Day 35 | Day 189 | Day 217 |  |  |  |
| GMT<br>(95% CI) | 40.0<br>(27.0, 59.5) | 10.9<br>(9.12, 13.0) | 265.3<br>(192.9, 364.7) | 14.5<br>(11.2, 18.7) | 10.66<br>(9.52, 11.93) | 214<br>(140.2, 326.8) |  |  |  |
| GMFR Day 217:Day 35<br>(95% CI) | 6.61<br>(4.3, 10.1) |  |  | 14.8<br>(7.74, 21.37) |  |  |  |  |  |
| GMFR Day 217:Day189<br>(95% CI) | 24.4<br>(16.6, 35.7) |  |  | 20.1<br>(10.56, 29.25) |  |  |  |  |  |

CI = confidence interval; GMFR = geometric mean fold rise; GMT = geometric mean titer; hACE2 = human angiotensin-converting enzyme 2; IC<sub>50</sub> = 50% inhibition concentration; SARS-CoV-2 = severe acute respiratory syndrome coronavirus 2
Note: Assay methodology is fit-for-purpose.

**Table 5.** Anti-rS IgG Geometric Mean Titers after Primary and Booster Vaccination for the Ancestral and Variant SARS-CoV-2 Strains by Study Day for Participants Receiving NVX-CoV2373.

| Parameter | Anti-rS IgG Activity (EC<sub>50</sub>) |  |  |  |  |  |  |  |  |
| --- | --- | --- | --- | --- | --- | --- | --- | --- | --- |
| Strain | Ancestral |  |  | Alpha |  |  | Beta |  |  |
| Study Day | Day 35 | Day 189 | Day 217 | Day 35 | Day 189 | Day 217 | Day 35 | Day 189 | Day 217 |
| GMT<br>(95% CI) | 60,742<br>(42,176, 87,481) | 5,361<br>(3,782, 7,599) | 327,758<br>(225,862, 475,623) | 24,333<br>(15,234, 38,865) | 2,739<br>(1,777, 4,223) | 235,145<br>(152,897, 361,636) | 40,416<br>(28,091, 58,147) | 4,066<br>(2,767, 5,975) | 264,321<br>(177,965, 392,582) |
| GMFR Day 217:Day 35<br>(95% CI) | 5.4<br>(3.3, 8.7) |  |  | 9.7<br>(5.6, 11.9) |  |  | 6.5<br>(4.0, 10.8) |  |  |
| GMFR Day 217:Day 189<br>(95% CI) | 61.2<br>(38.9, 96.4) |  |  | 85.9<br>(50.4, 146.1) |  |  | 65.0<br>(40.0, 105.4) |  |  |
| Strain | Delta |  |  | Omicron |  |  |  |  |  |
| Study Day | Day 35 | Day 189 | Day 217 | Day 35 | Day 189 | Day 217 |  |  |  |
| GMT<br>(95% CI) | 26,097<br>(17,501, 38,916) | 3,143<br>(1,952, 5,059) | 290,782<br>(195,349, 432,836) | 11,119<br>(7,668, 16,121) | 1,413<br>(805.2, 2,481) | 103,800<br>(67,398, 159,860) |  |  |  |
| GMFR Day 217:Day 35<br>(95% CI) | 11.1<br>(6.5, 19.1) |  |  | 9.3<br>(8.8, 9.9) |  |  |  |  |  |
| GMFR Day 217:Day 189<br>(95% CI) | 92.5<br>(52.8, 162.4) |  |  | 73.5<br>(38.5, 140.2) |  |  |  |  |  |

Anti-rS IgG = anti-recombinant spike immunoglobulin G antibody; CI = confidence interval; EC<sub>50</sub> = 50% effective concentration; GMFR = geometric mean fold rise; GMT = geometric mean titer; SARS-CoV-2 = severe acute respiratory syndrome coronavirus 2

**Figure 5.**
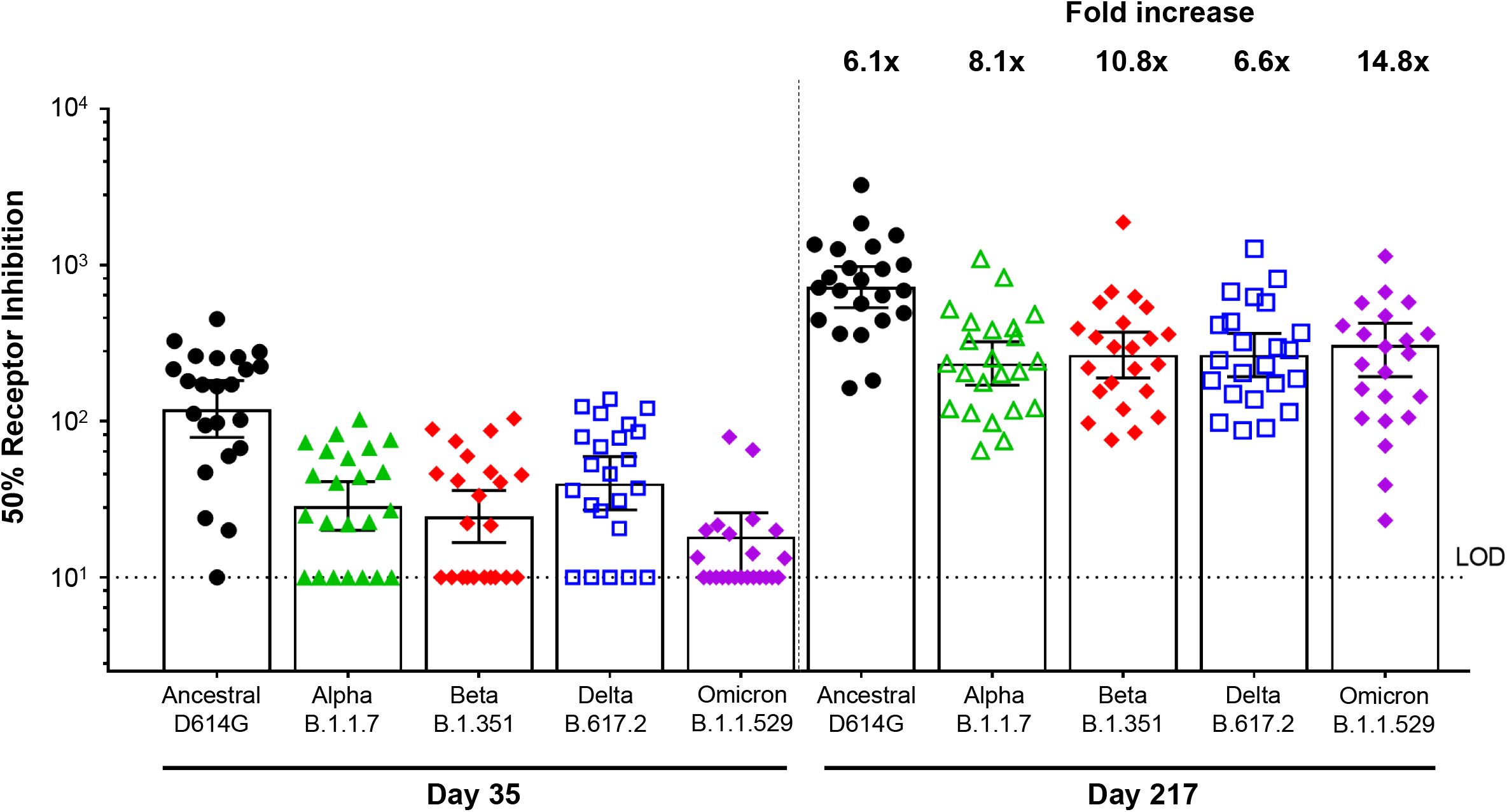
hACE2 Receptor Binding Inhibition Titers for SARS-CoV-2 Variants Post Primary Vaccination at Day 35 and Post-Booster at Day 217. hACE2 = human angiotensin-converting enzyme 2; LOD = limit of detection; SARS-CoV-2 = severe acute respiratory syndrome coronavirus 2 Note: Assay methodology is fit-for-purpose.

**Figure 6.**
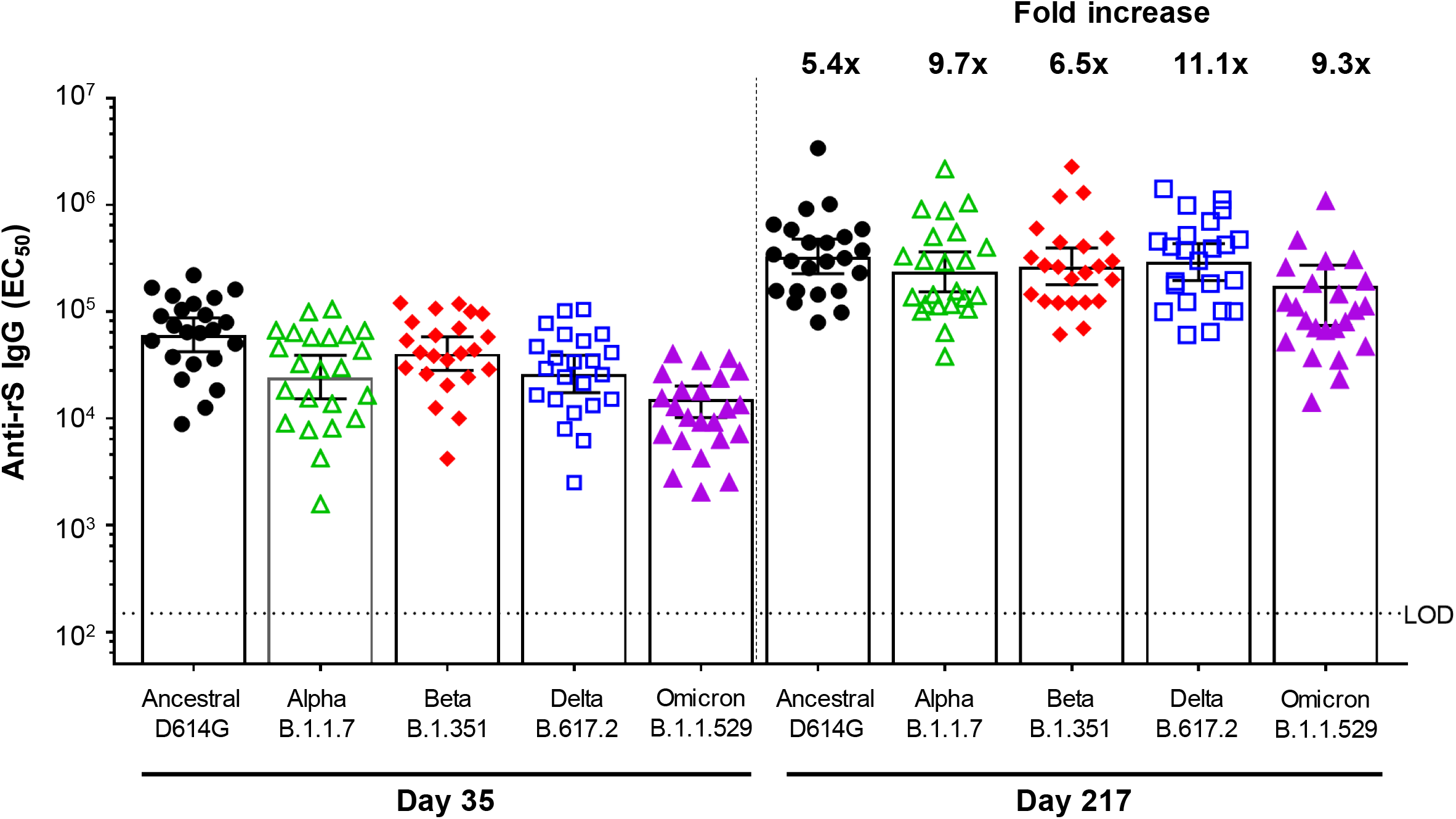
Anti-rS IgG Titers for SARS-CoV-2 Variants Post Primary Vaccination at Day 35 and Post-Booster at Day 217. Anti-rS IgG = anti-recombinant spike immunoglobulin G antibody; EC_50_ = 50% effective concentration; LOD = limit of detection; SARS-CoV-2 = severe acute respiratory syndrome coronavirus 2 Note: Assay methodology is fit-for-purpose.

## DISCUSSION

In this report, we describe the first available safety and immunogenicity data for a booster dose of NVX-CoV2373 in the context of an ongoing phase 2, randomized, observer-blinded, placebo-controlled study. Administration of a single booster dose of the vaccine approximately 6 months following the primary two-dose series resulted in an incremental increase in reactogenicity events along with significantly enhanced immunogenicity.

Prior to boosting at Day 189, anti-SARS-CoV-2 antibody titers in immunized participants were markedly lower when compared with samples taken after the primary vaccination series at Day 35 (Group B IgG and MN_50_ GMTs lowered from 43,905 EU to 6,064 EU and 1,470 to 63, respectively). Correlating falling antibody titers in individuals immunized or previously exposed to COVID-19 with waning immunity can be complex, though there is established evidence that the presence of neutralizing antibodies are strongly indicative of protection against symptomatic COVID-19. Further, vaccines that generate higher neutralizing antibody titers (NVX-CoV2373, mRNA-1273, BNT162b2, and Sputnik V) have demonstrated higher VE in clinical trials than those associated with lower titers (ChAdOx1 nCoV-19, Ad26.COV2.S, and CoronaVac) (Hamady 2021, Khoury 2021).

Maintaining robust immunity may be especially important in the presence of SARS-CoV-2 variants that have shown to be a cause for breakthrough infections. Other mechanisms for maintaining long-term immunity among multiple variants may involve “vaccine mixing” with heterologous boosters or utilizing vaccine technologies that result in broadly cross-reacting immunity, as observed with an adjuvanted recombinant nanoparticle influenza vaccine (Hamady 2021, Shinde 2020).

In the present study, antibody responses to the booster were assessed for the ancestral vaccine strain as well as for more recent SARS-CoV-2 variants including Alpha, Beta, and Delta. For the ancestral strain, IgG titers at Day 217 were approximately 34-fold higher than the pre-booster Day 189 titers while neutralizing antibody titers increased approximately 96-fold after the booster. Both IgG and MN titers after the booster were > 4-fold higher than those seen after the primary two-dose series at Day 35, which is notable as the Day 35 titers corresponded to high levels of clinical efficacy in both a UK phase 3 study (89.7%) as well as in a USA/Mexico phase 3 study (90.4%) (Dunkle 2021, Heath 2021). When broken down by age group, higher fold increases were seen for older adults (≥ 60 to ≤ 84 years of age) compared to younger adults (≥ 18 to ≤ 59 years of age). This finding suggests that a booster dose may have added benefit in older adults as their antibody responses following the primary two-dose vaccination series were lower than those seen in younger adults.

For the Beta variant, 40-to 50-fold increases in IgG and MN antibody titers were seen following the booster and IgG titers were approximately 4-fold higher than those seen for the ancestral strain after the primary vaccination series. Unlike the observation with IgG, MN_50_ GMTs for the Beta variant were lower following the booster than those for the ancestral strain following the primary vaccination series (GMT 661 vs 1,470) in alignment with the known decreased neutralizing responses for this variant. While the worldwide prevalence of Beta has recently decreased to <1% from a high of 8% in April 2021, antibody responses to this variant remain of interest due to the E484K mutation found in this variant. E484K has been found responsible for marked decreases in neutralization titers for vaccines, monoclonal antibodies, and convalescent sera, and the mutation remains in the P.1, P.2, and the Mu variants of SARS-CoV-2 (Nextstrain 2021, Harvey 2021).

For the Delta and Omicron variants of SARS-CoV-2, which as of December 2021 represent nearly 100% of all strains circulating, 24.4-fold (Delta) and 20.1-fold (Omicron) increases in functional hACE2 receptor binding inhibition titers were seen when comparing the post-booster titers from Day 217 to Day 189 titers (Nextstrain 2021). Anti-rS IgG activity compared at these same time points found 92.5-fold (Delta) and 73.5-fold (Omicron) higher titers associated with the booster.

These findings of significantly increased antibody titers following boosting are important as they come during a time when SARS-CoV-2 vaccine booster doses are being widely considered or implemented by a number of countries in order to counteract the waning antibody titers and decreased effectiveness for approved vaccines (Hamady 2021). The continued high levels of SARS-CoV-2 circulation combined with increasing immunological pressure may also give rise to new escape variants; increasing individuals’ titers to these variants through use of a booster dose may extend protection in previously vaccinated individuals and combat the spread of SARS-CoV-2 variants.

The incidence of both local and systemic reactogenicity was higher following the 6-month booster dose compared to the previous doses reflecting the increased immunogenicity seen with the third dose. However, the incidence of Grade 3 or higher events remained relatively low with only fatigue (12.2%) being recorded by greater 10% of participants. In total, five Grade 4 (potentially life threatening) solicited local and systemic adverse events were reported. All five of these events (pain, tenderness, headache, malaise, and muscle pain) were reported by the same participant in the active booster group concurrently with an adverse event of drug hypersensitivity related to the vaccine. The drug hypersensitivity event was assessed as mild in severity. The participant did not seek any medical attention for this event, and all the participant’s symptoms resolved over a period of 6 days.

With the caveat that the data are drawn from different studies, it appears that the reactogenicity rates seen after a third dose of NVX-CoV2373 are generally similar to those seen with a booster dose of the Pfizer/BioNtech and Moderna vaccines and elevated compared to the Oxford/AstraZeneca vaccine (Flaxman 2021, Choi 2021, FDA 2021). While the incidence of unsolicited adverse events following the booster was higher in vaccine recipients (12.4%) compared to placebo recipients (11.0%), all events were classified as mild or moderate in severity. Medically attended adverse events, PIMMCs, and SAEs occurred infrequently following the booster dose and were balanced between vaccine and placebo groups.

Our study was subject to certain limitations. As these results are from an ongoing phase 2 study conducted with a limited sample size, clinical efficacy of the booster dose was not evaluated. In addition, between-strain comparisons of antibody data should be made with caution, as correlations with clinical efficacy may vary for each variant of COVID-19.

Overall, a single booster dose of NVX-CoV2373 administered approximately 6 months after the primary series induced a substantial increase in humoral antibodies that was > 4-fold higher than antibody titers associated with high levels of efficacy in two phase 3 studies while also displaying an acceptable safety profile. These findings support use of the vaccine in booster programs.

## Data Availability

Data Availability
Data Sharing Statement will be available with the full text of this article upon publication.

https://clinicaltrials.gov/ct2/show/NCT04368988

## Acknowledgments

We thank all of the study participants who volunteered for this study. Graphics assistance for this manuscript was provided by Ashfield MedComms, supported by Novavax, Inc.

